# Palette polygenic risk score framework improves risk prediction by capturing clinical heterogeneity of type 2 diabetes

**DOI:** 10.64898/2026.01.12.25342123

**Authors:** Akimitsu Miyake, Hayato Tanabe, Akira Narita, Takafumi Ojima, Tomoki Kyosaka, Chinatsu Gocho, Rieko Sakurai, Jun Takayama, Hajime Yamakage, Kenichi Tanaka, Junichiro James Kazama, Noriko Satoh-Asahara, Michio Shimabukuro, Gen Tamiya

## Abstract

Polygenic risk scores (PRSs) are typically constructed under the assumption of a single, homogeneous disease phenotype. However, many common diseases exhibit considerable clinical heterogeneity and encompass multiple subtypes with distinct etiologies and clinical characteristics. As a result, conventional PRSs often overlook differences in underlying biological pathways among disease subtypes, consequently limiting predictive accuracy and cross-ancestry transferability. To address this challenge, we propose the “palette PRS,” a framework that integrates a set of partitioned polygenic scores (pPSs) for biologically interpretable pathways with subtype-specific weights. This approach can flexibly capture the relative contributions of multiple pathways within each individual and provides a unified risk score. We applied this framework to type 2 diabetes (T2D), a clinically highly heterogeneous disease. For T2D, previous machine learning-based studies have identified four distinct subtypes and 12 biologically interpretable pathways derived from 650 genome-wide significant variants. Building on these established findings, we employed an elastic net model incorporating subtype membership probabilities to derive subtype-optimized palette PRS through the weighted integration of the pPSs of these 12 pathways. Our palette PRS showed superior predictive performance, with particularly high accuracy for the severe insulin-deficient diabetes (SIDD) subtype (AUC=0.744), compared with both conventional T2D PRS (AUC = 0.661) or subtype-stratified GWAS-based PRS (AUC = 0.547). Moreover, our palette PRS exhibited substantial cross-ancestry transferability between East Asian and European populations. This strategy represents a major step toward clinically actionable, subtype-optimized risk prediction and personalized prevention in T2D worldwide.

## Introduction

The genetic risk of common diseases arises from the cumulative effects of numerous variants across the genome. Polygenic risk scores (PRSs) quantify this genetic risk, enabling stratification of individuals by the risk and supporting precision prevention strategies [1,2,3,4]. In recent years, the expansion of large-scale genome-wide association studies (GWAS), particularly those including populations of diverse ancestry, has further improved PRS performance [5,6]. However, their clinical utility remains limited, mainly due to challenges in modest predictive accuracy and poor cross-ancestry transferability [7,8]. One of the key limitations of most GWAS and PRS frameworks is their reliance on binary case-control definitions, which collapse clinically heterogeneous cases into a large single category and thereby obscure clinical heterogeneity and underlying pathophysiological mechanisms [9].

Type 2 diabetes (T2D) is a representative example of this limitation [9]. Although multi-ancestry GWAS has expanded sample sizes and modestly improved overall T2D risk prediction [10,11], conventional PRS still treats T2D as a single disease entity despite its now well-recognized clinical heterogeneity [12–20], resulting in relatively low predictive performance (AUC∼0.60–0.65). Among existing subtype classifications, Ahlqvist’s k-means clustering analysis applied to a few clinical measurements in a large cohort has most reproducibly identified four distinct T2D subtypes with differing clinical and genetic profiles: severe insulin-deficient diabetes (SIDD), severe insulin-resistant diabetes (SIRD), mild obesity-related diabetes (MOD), and mild age-related diabetes (MARD) [12,21]. Accounting for clinical heterogeneity across subtypes into risk prediction holds promise for improving performance, as shown in subtype-optimized risk models for breast cancer [22]. The most straightforward approach would be to construct PRS directly from subtype-stratified GWAS. In practice, though, this strategy is statistically inefficient, as it inevitably reduces the sample size for each subtype, thereby worsening discovery power and downstream predictive accuracy.

To overcome this obstacle, we utilized biologically interpretable pathways underlying clinical heterogeneity and the differences in their relative importance across the machine-learned subtypes [23,24]. A biologically interpretable pathway corresponds to a cluster of T2D-associated variants identified by grouping variants that share similar multi-trait association patterns across T2D-related traits [25]. Several studies have attempted to group T2D-associated variants into a small number of biologically interpretable clusters [25–27]. Most recently, Smith et al. successfully identified 12 clusters using Bayesian non-negative matrix factorization (bNMF) with 650 T2D genome-wide significant variants and GWAS summary statistics for 110 T2D-related traits [27], which can be biologically annotated as 12 pathways. For each identified pathway, an individual’s genetic susceptibility can be summarized as a set of partitioned polygenic scores (pPSs). Recent studies show that pPSs are useful for explaining biological heterogeneity and are being validated across populations [27–31]. While each pPS offers meaningful biological insight, previous study indicates that the effect of each individual pPS on T2D risk is predicted to be small in magnitude [32]. Given that T2D is thought to be driven by the combined influence of multiple biological pathways rather than any single pathway [33,34], these findings highlight opportunities to further improve prediction by integrating signals across pathways.

Here, we propose the palette PRS, a framework that integrates an individual’s set of pPSs with subtype-specific pathway weights to generate a unified risk score (Fig.1a). This framework is a predictive approach inspired by the McCarthy’s “palette model” hypothesis [33], where a mixture of unrevealed biological pathways may shape a multifactorial and continuous spectrum of T2D predisposition. Using 650 genome-wide significant variants from multi-ancestry cohorts [27], we employed an elastic net model incorporating subtype membership probabilities to generate subtype-optimized risk scores by weighted integration of 12 biologically informed pPSs (Materials & Methods). Using large-scale T2D datasets, we evaluated whether the palette PRS could improve T2D risk prediction compared with conventional PRSs.

**Fig.1:**
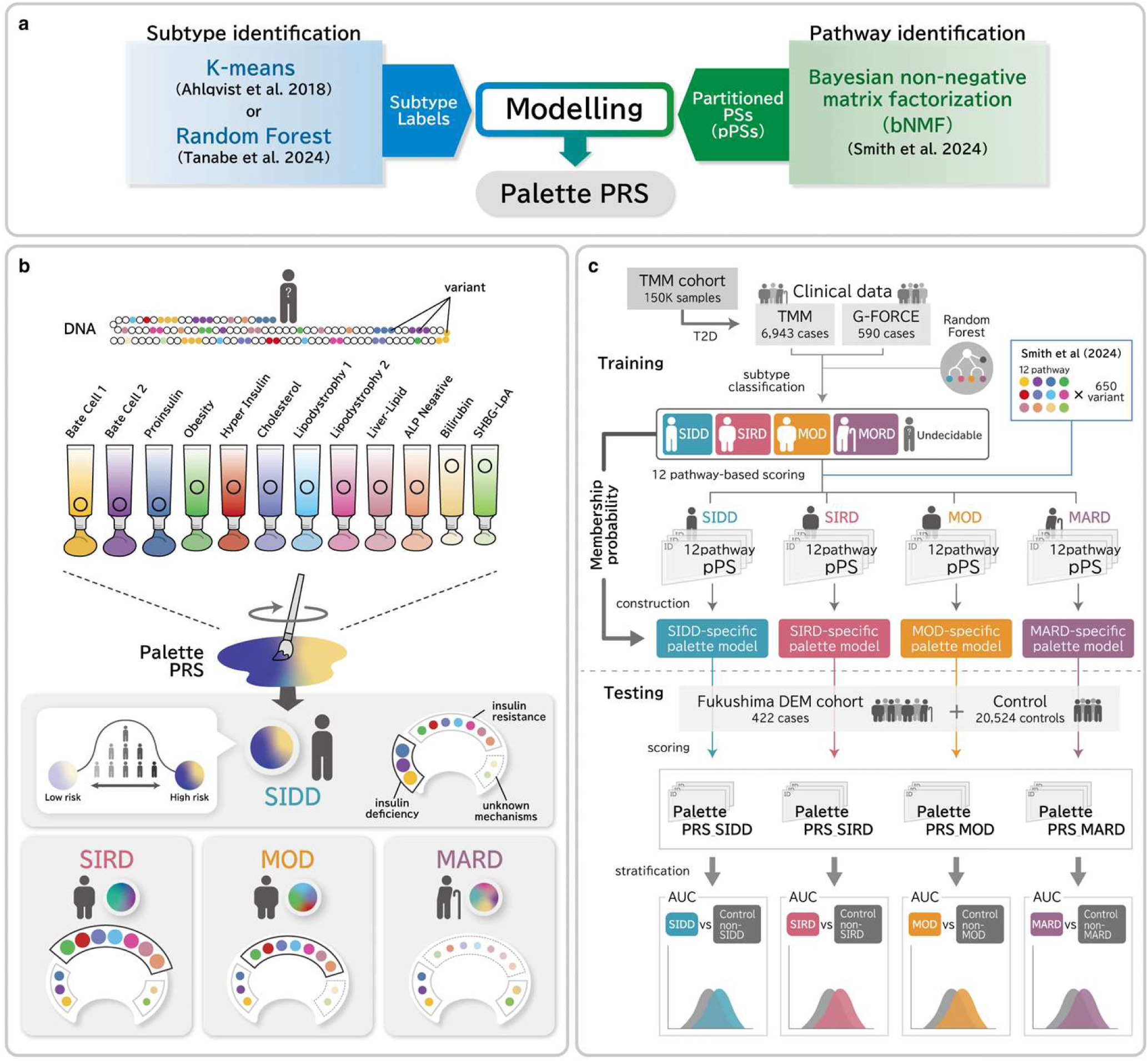
Conceptual overview of the palette PRS framework and study design. **a.** Construction of the palette PRS from subtype labels and pPSs. Subtypes are conventionally identified by k-means clustering of Ahlqvist’s variables (age at diagnosis, BMI, HbA1c, HOMA2-B, and HOMA2-IR) [12]. However, when some of these variables are missing, as is the case for most biobank datasets used in this study [9], our random forest classifier provides an effective alternative because it handles incomplete data more robustly [35]. pPSs are derived from biologically interpretable pathways defined by bNMF, and individual-level scores are computed for each pathway. These subtype labels and pPSs are integrated via Elastic Net model to produce the palette PRS. **b.** Conceptual architecture of the palette polygenic risk prediction. Each pathway is treated as a distinct “primary color,” with its pPS interpreted as the saturation of that color. Mixture coefficients (“paint amounts”) that capture subtype-specific pathway contributions are estimated from clinical subtype information. The final palette PRS is expressed as a weighted composition of three elements: (1) pathway (“primary color”), (2) pPS magnitude (“saturation”), and (3) subtype-specific pathway weights (“paint amounts”). **c.** Overview of model development and evaluation for the palette PRS. In each cohort, we first classified individuals with T2D into subtypes using a random forest classifier trained on clinical variables, retaining subtype membership probabilities to account for uncertainty in subtype assignment. For individuals whose maximum subtype assignment probability was less than 0.5, we designated the subtype as ‘Undecidable’ to avoid forcing uncertain labels into downstream modeling [35]. We then computed 12 pPSs in the TMM and G-FORCE training cohorts using pathway weights derived from the bNMF analysis of 650 multi-ancestry T2D-associated variants [27] and constructed subtype-optimized prediction models that incorporated these subtype membership probabilities. We used control samples (*n_control_* = 41,049) consisting of non-diabetic individuals from TMM and randomly split them into two equal subsets for independent use in model development (training) and external validation (testing). Finally, we applied each subtype-specific model to the external validation cohort, Fukushima DEM, to evaluate the predictive performance of the palette PRS.

## Results

### Study overview

The palette PRS framework integrates subtype-label information with a set of pPSs (Fig. 1a). Subtype labels are identified either by k-means clustering [12] or by our random forest classifier trained to predict them from routinely available clinical data [35]. In parallel, pPSs are derived from biologically interpretable pathways obtained via bNMF [25–27], and individual-level pPSs are computed for each pathway. An elastic net model then integrates the subtype label information with the pPSs to estimate the palette PRS (Fig.1a).

As schematically shown in Fig. 1b, we conceptualize the palette PRS framework as a model in which 12 pPSs, each derived from 650 T2D-associated variants, serve as the 12 “primary colors.” In this schematic explanation, each color labels a pathophysiological pathway; the saturation corresponds to the magnitude of the pPS for that pathway, and the proportion of each color in the mixture reflects its contribution to the final subtype-optimized composite score (Fig. 1b).

Using our random forest classifier [35] similar to the Ahlqvist’s k-means application, we classified participants with T2D from the Tohoku Medical Megabank (TMM) (*n*_T2D_ = 6,943) and G-FORCE (*n*_T2D_ = 590) cohorts into four major subtypes (SIDD, SIRD, MOD, and MARD), while borderline or classification-ambiguous cases were categorized as an additional ‘Undecidable’ group. Each subtype exhibited baseline characteristics consistent with its previously reported clinical features (Supplementary Figs. 1–3; Supplementary Tables 1 and 2) (Fig.1c). We subsequently calculated each pPS of the pathways for all participants and estimated subtype-specific pathway weights through elastic net model training (Materials & Methods, Supplementary Tables 3). We trained the model using participants from the TMM and G-FORCE cohorts and conducted external validation in the Fukushima DEM cohort of individuals with *T2D (n*_T2D_ *= 422)*. We evaluated predictive performance for each target subtype against a combined reference group comprising the remaining T2D subtypes, ‘Undecidable’ cases, and non-diabetic controls (*n*_control_ = 20,524) (Fig.1c).

### PRS prediction of T2D subtypes and overall T2D

For each subtype, we evaluated discriminative performance using the area under the receiver operating characteristic curve (AUC) in a one-vs-rest approach (e.g., SIDD cases vs. non-SIDD and controls) (Table 1). The palette PRS demonstrated the highest performance for SIDD (AUC = 0.744 [95% CI: 0.668–0.819]), followed by MOD (AUC = 0.680 [95% CI: 0.628–0.732]), MARD (AUC = 0.649 [95% CI: 0.608–0.690]), SIRD (AUC = 0.648 [95% CI: 0.585–0.711]). By contrast, the PRSs derived from subtype-stratified GWAS using PRS-CS-auto [37] showed limited predictive accuracy across all subtypes (SIDD: AUC = 0.547 [95% CI: 0.467–0.627], SIRD: AUC = 0.509 [95% CI: 0.481–0.580], MOD: AUC = 0.549 [95% CI: 0.493–0.605], MARD: AUC = 0.554 [95% CI: 0.510–0.598]) and were consistently outperformed by the palette PRS (Table 1).

**Table 1.** Comparative predictive performance of palette PRS and stratified GWAS-based PRS in T2D subtypes.

| Population with<br>T2D subtypes | AUC (+95%CI) in testing dataset |  |
| --- | --- | --- |
|  | Palette PRSs | Subtype-stratified GWAS PRSs |
| SIDD | 0.744 (0.668–0.819) | 0.547 (0.467–0.627) |
| SIRD | 0.648 (0.585–0.711) | 0.509 (0.481–0.580) |
| MOD | 0.680 (0.628–0.732) | 0.549 (0.493–0.605) |
| MARD | 0.649 (0.608–0.690) | 0.554 (0.510–0.598) |

Next, we evaluated a conventional T2D PRS generated by summing the 650 genome-wide significant variants weighted by their β coefficients from the multi-ancestry GWAS meta-analysis (MVP and DIAMANTE) [36] (Table 1). As expected, the conventional PRS achieved limited discrimination (AUC = 0.661[95% CI: 0.634–0.687]), falling short of the subtype-optimized palette PRS (Table 1).

We then examined the discriminative performance of individual pPSs within each subtype and for overall T2D (Fig. 2; Supplementary Table 4). For overall T2D, the pPS of “Beta Cell 1” achieved an AUC of 0.632 [0.604–0.660]. When stratified by subtype, however, the “Beta Cell 1” pPS consistently showed the highest performance among the 12 pPSs across all subtypes, with the strongest result observed in SIDD (AUC = 0.691, 95% CI 0.620–0.762), followed by MOD, MARD, and SIRD showing AUCs of 0.636 [0.583–0.689], 0.623 [0.580–0.663], and 0.593 [0.525–0.662], respectively. None of the 12 pPSs, including “Beta Cell 1”, exceeded the performance of their corresponding palette PRSs within any subtype.

**Fig.2:**
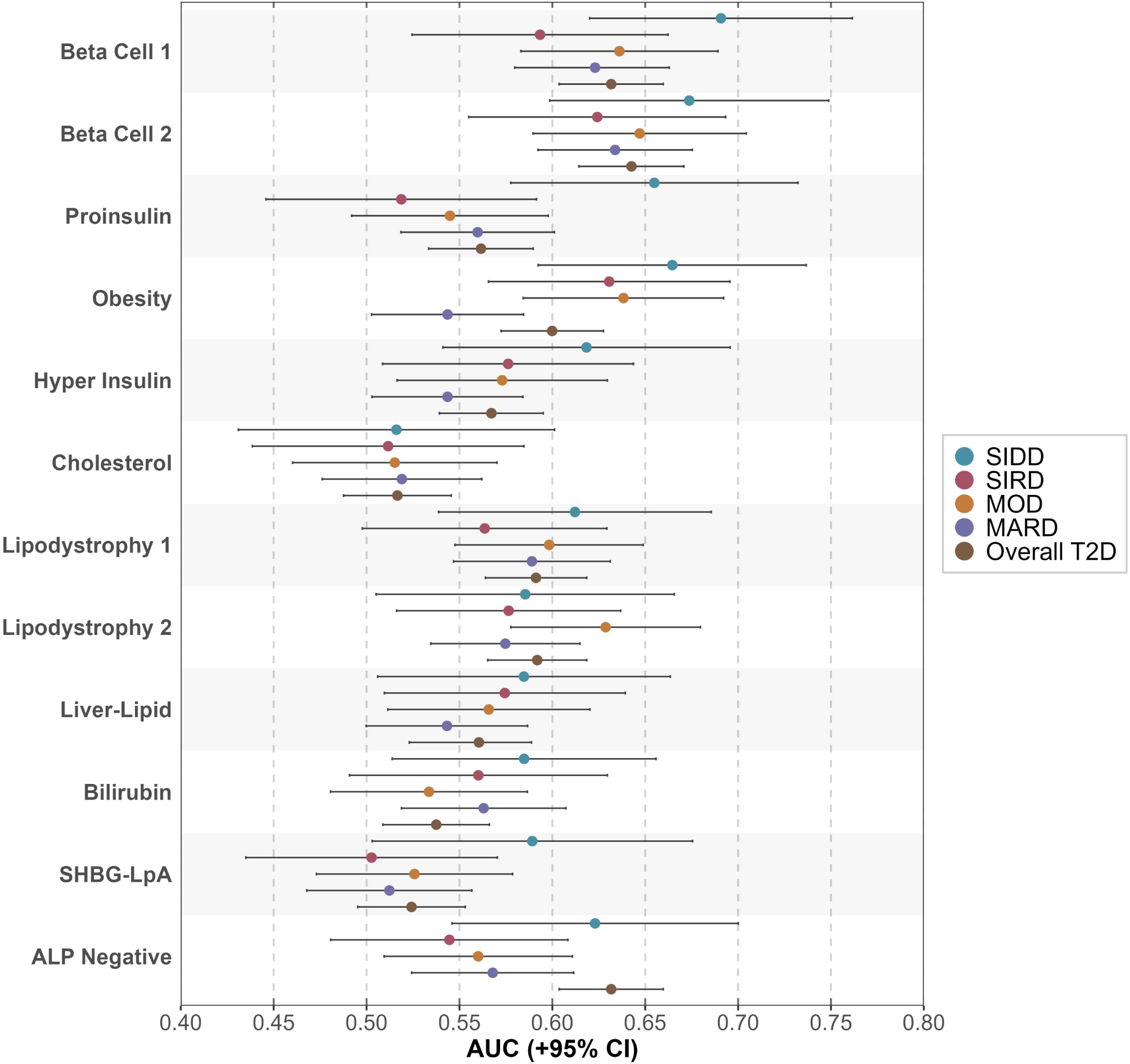
Predictive performance of pPS in T2D. Predictive accuracy of individual pPSs for each subtype and overall T2D. Each pPS is derived from one of the 12 biologically interpretable pathways. All polygenic risk models were evaluated without covariate adjustment. Exact values are provided in Supplementary Table 4.

### Subtype-specific risk stratification by palette PRS

Compared with the reference group (4th PRS decile, 30–70th percentile), individuals in the top decile (90–100th percentile) showed significantly higher odds for each subtype: SIDD, OR=5.6 (95% CI: 3.1–9.9; Fisher’s exact test *P* =1.33 x 10^-7^); SIRD, OR=2.3 (95% CI: 1.3–4.2; *P*=7.00 x 10^-3^); MOD, OR=3.4 (95% CI: 2.2–5.3; *P* =3.42 x 10^-7^); and MARD, OR=3.0 (95% CI: 2.1–4.3; *P* =9.03 x 10^-9^) (Fig. 3a,b; Supplementary Table 5). Among subtypes, SIDD had the highest odds in the top PRS decile, indicating the most effective risk stratification. Calibration analysis showed overall agreement between predicted and observed risks (Supplementary Fig. 4).

**Fig.3:**
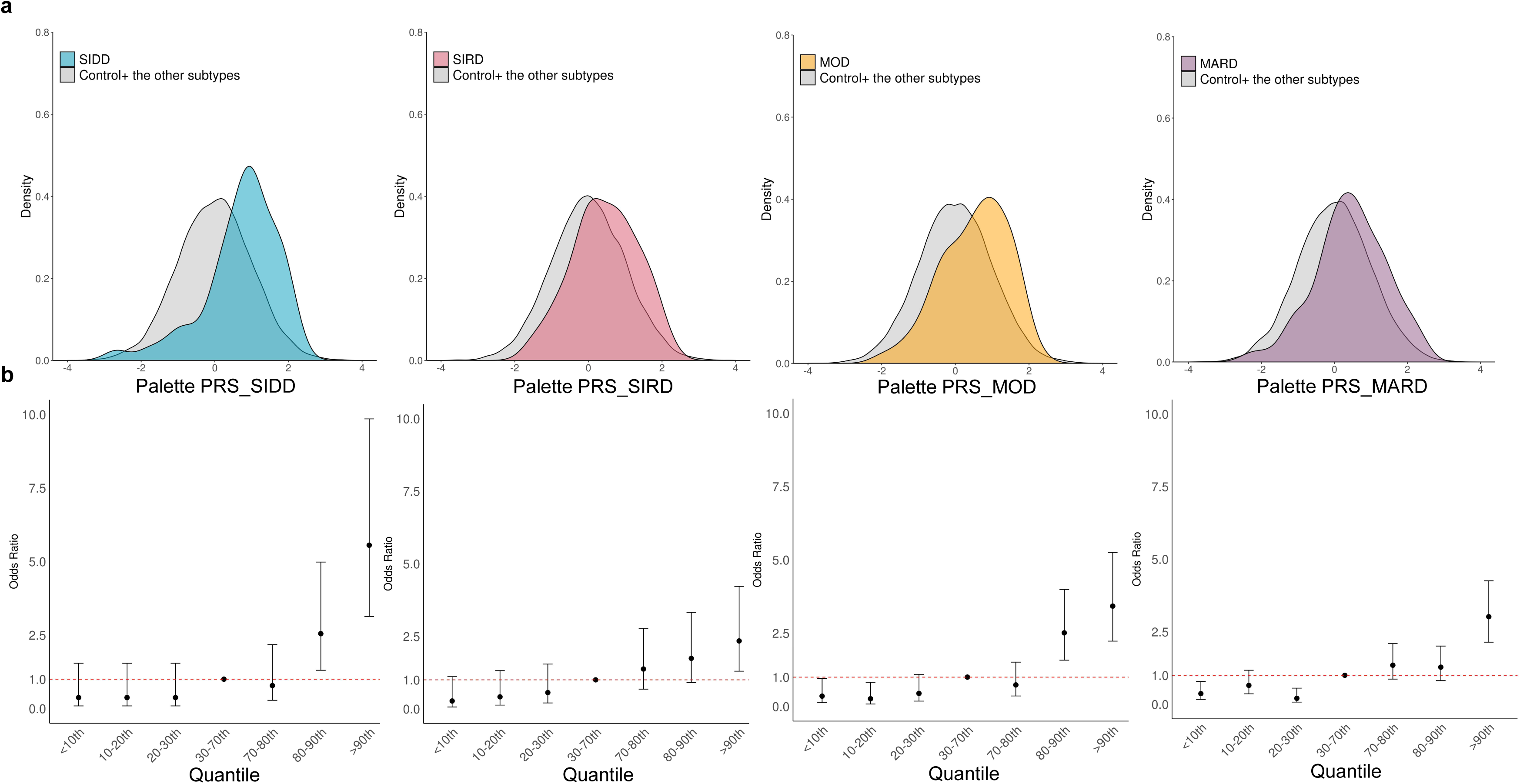
Palette PRS distribution and odds ratios by quantile. a. Density distributions of the normalized palette PRS for each T2D subtype: SIDD (blue), SIRD (red), MOD (orange), and MARD (purple). Gray denotes controls and samples from the other subtypes. b. Odds ratios for each subtype across palette PRS deciles. Participants were divided into seven groups by palette PRS, with the 30–70th percentile serving as the reference group (red dashed line). Error bars indicate 95% confidence intervals.

## Discussion

In this study, we proposed the palette PRS, a polygenic prediction framework optimized for T2D subtypes, and evaluated its predictive performance. The palette PRS outperformed both a conventional overall T2D PRS and subtype-stratified GWAS-based PRSs, with the largest improvement observed for SIDD, followed by MOD. Although “Beta Cell 1” was the strongest individual pPS for SIDD, it still did not exceed the performance of the corresponding palette PRS. These results are consistent with previously reported high heritability for SIDD and MOD [21], supporting the utility of this approach for subtype-optimized prediction.

Our palette PRS is built upon previously defined pPSs, which quantify genetic susceptibility attributable to specific biological pathways. Although all pPSs originate from the same set of 650 variants, bNMF assigns a weight of zero to pathway-specific subsets of variants, with each pathway receiving a different subset of zero-weighted variants [27]. Our model assigns subtype-specific weights to these pPSs, capturing both shared and subtype-specific genetic components and optimizing pathway-specific contributions to genetic risk within each subtype. Instead of re-discovering subtype-specific variants on smaller subtype-stratified samples, the palette PRS uses a common foundation of 650 established T2D-associated variants identified in a large multi-ancestry GWAS. Clinical heterogeneity across subtypes is captured not by splitting the dataset, but by optimizing pathway weights for each subtype. This approach avoids the loss of statistical power and enables subtype-optimized risk prediction that reflects distinct biological effects. Such an advantage is particularly important for uncommon and severe subtypes such as SIDD, where limited case numbers often hinder the reliable estimation of genetic effects.

The superior performance of the palette PRS stems from its distinct way of modeling clinical and biological heterogeneity among T2D subtypes, which differs radically from conventional PRS approaches. Conventional, overall PRSs that treat T2D as a single homogeneous disease are affected by differences in subtype composition across cohorts and tend to reflect predominantly the larger subtype within each cohort. For example, a cohort enriched for obesity-related cases, such as G-FORCE, may yield an overall PRS overrepresenting SIRD or MOD components. On the other hand, subtype-stratified GWAS-based PRSs, while accounting for clinical heterogeneity, lose statistical predictive power due to reduced sample sizes. Our palette PRS overcomes both limitations by flexible modeling of the relative contributions of biological pathways across subtypes. By optimizing pathway weights separately for each subtype, the palette PRS gets around the dilution of genetic signals caused by mixing effects of opposite directions across subtypes in overall PRSs and prevents power loss by using a shared foundation of T2D-associated variants derived from large-scale multi-ancestry GWAS, rather than conducting separate underpowered GWAS for each subtype.

As an additional analysis, we evaluated the transferability of the palette PRS, testing whether models trained in a population of one ancestry could be applicable to populations of another ancestry (Fig. 4). Intriguingly, a model trained in the UK Biobank (European ancestry) [38] performed comparably when validated in Japanese Fukushima DEM cohort. For SIDD, the AUC was 0.741 [95% CI: 0.666–0.816]; for SIRD, 0.639 [95% CI: 0.574–0.704]; for MOD, 0.699 [95% CI: 0.647–0.750]; and for MARD, 0.650 [95% CI: 0.610–0.690]. These values were nearly identical to those obtained using models trained in East Asian data. The improved transferability of the palette PRS can be attributed to several key differences from conventional PRSs: they aggregate high-dimensional variant information into a single summary score, making them sensitive to ancestry-related differences in linkage disequilibrium and allele frequencies [39]. In contrast, the palette PRS utilizes a curated set of 650 T2D-associated variants identified across a multiple ancestral GWAS [27], capturing shared genetic signals from diverse populations rather than relying on any single ancestry. Within the framework of our palette PRS, variant effects are integrated across 12 biologically annotated pathways using bNMF-derived weights to produce pPSs that summarize the collective genetic contribution of related variants. Importantly, because these pPSs are calculated using the same set of weights derived from a single bNMF, cross-ancestry transferability depends only on the 12 pathway-level weights, which likely contributes to the model’s stable performance across ancestries. Moreover, subtype-specific weighting allows the model to remain robust even when a subtype composition differs across populations, mitigating the bias described earlier.

**Fig.4:**
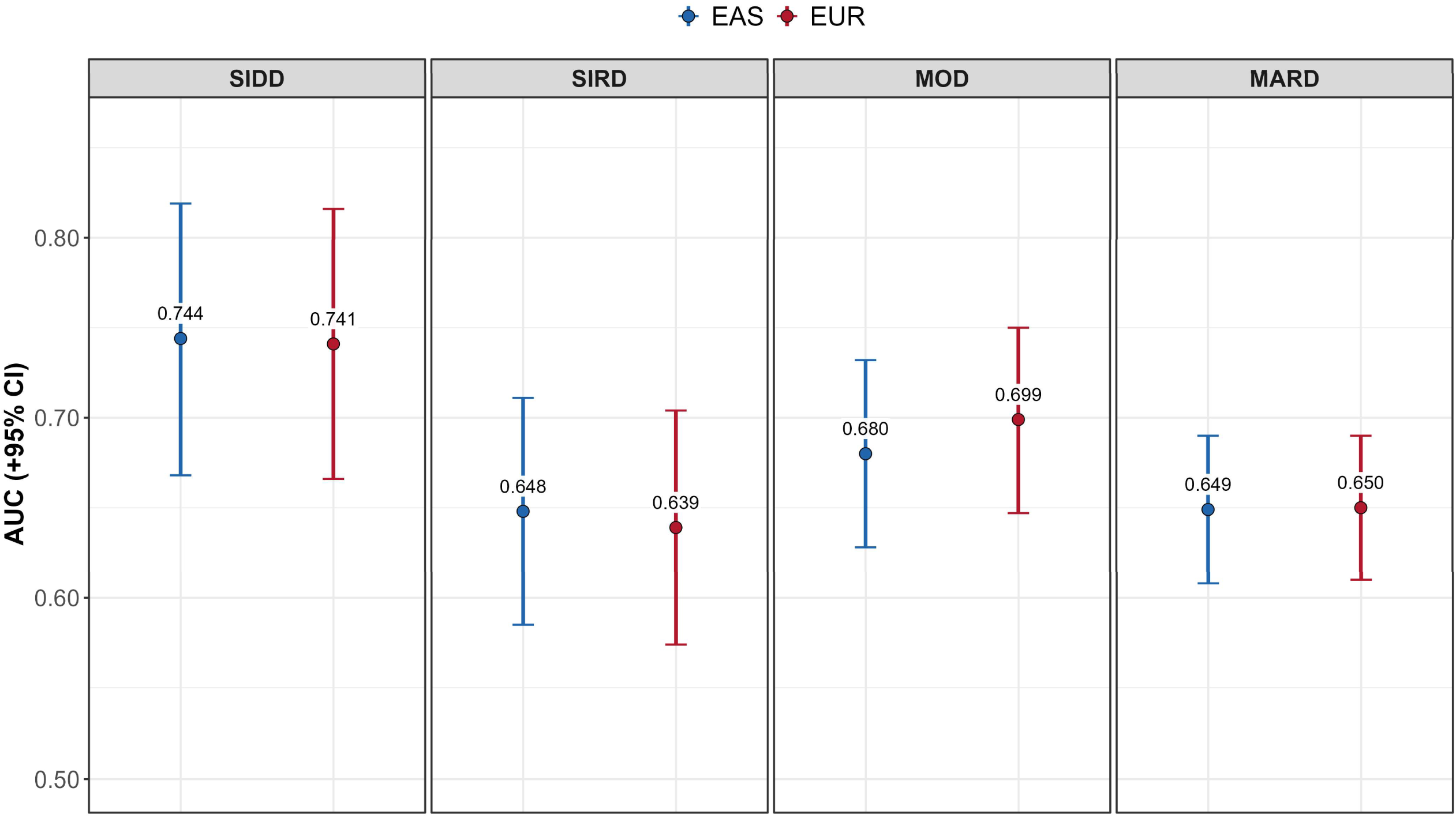
Prediction accuracy of the palette PRS trained on UK Biobank British individuals. Discriminative performance of the palette PRS. The UK Biobank dataset (*n_T_*_2*D*_ = 18,705; *n_control_* = 413,815) was used to assign T2D subtypes via random forest classification (Supplementary Figs. 6, 7; Supplementary Table 1) and to train subtype-specific palette PRS models (Supplementary Table 3). Predictive performance was evaluated in the Fukushima DEM cohort.

From a clinical perspective, our palette PRS enables risk prediction that offers clinically actionable insights informed by key features of each subtype, thereby moving beyond conventional one-size-fits-all strategies. Effective diabetes prevention requires accurate recognition of clinical heterogeneity of the disease and integration of genetic, clinical, and environmental information into precision prevention strategies [40]. By identifying individuals with high genetic risk for specific subtypes, the palette PRS potentially enables tailored prevention and management. For instance, β-cell–preserving approaches may benefit those at high risk of SIDD, while insulin resistance–focused care may suit those predisposed to SIRD [41]. Since subtypes differ in complication risk, early prediction can guide targeted monitoring. Moreover, because generic risk alerts often have limited behavioral impact [42], subtype-specific, actionable risk information may better drive effective interventions.

Our study has several limitations. First, in the TMM cohort, the absence of C-peptide required the use of estimated HOMA2 indices, introducing additional uncertainty that may have affected subtype classification. Although low-confidence individuals were assigned to an ‘Undecidable’ group [35], some misclassification may still have occurred. To overcome this limitation, large-scale biobank studies collecting more detailed phenotypic information, including C-peptide and age at onset, will be important for refining subtype classification and improving predictive performance. Second, the palette PRS was developed using the set of 650 variants incorporated at the bNMF stage, which may not capture the full spectrum of T2D-associated loci. Expanding the variant set by larger GWAS could further enhance predictive accuracy in future analyses. Third, our testing dataset was restricted to Japanese populations. Although transferability analysis using UK Biobank data suggested broader applicability, validation in additional ancestries, including South Asian, African, and admixed populations, will be needed to confirm generalizability. Finally, even within subtypes such as SIDD, substantial interindividual variability in genetic risk remains. Comparing complications and other clinical outcomes between individuals with high and low genetic risk within the same subtype will be an important direction for future research.

In conclusion, we proposed the palette PRS, a novel framework for polygenic risk prediction in T2D subtypes. This design achieved higher predictive accuracy than both conventional overall and subtype-stratified GWAS-based PRSs, with the greatest improvement observed for SIDD, and demonstrated stable performance across ancestries, suggesting advantages in cross-population transferability. Incorporating such subtype-specific genetic information may enable more personalized and effective prevention strategies. Future studies across diverse ancestries and clinical settings are warranted to confirm its generalizability and clinical utility. Beyond T2D, this framework could be applied to other complex diseases characterized by clinical and biological heterogeneity.

## Materials and Methods

### Training datasets

#### Tohoku Medical Megabank (TMM)

The Tohoku Medical Megabank (TMM) project is a large-scale prospective cohort in northeastern Japan, comprising the Community-Based Cohort Study (TMM CommCohort Study) and the Birth and Three-Generation Cohort Study (TMM BirThree Cohort Study) [43,44], totaling approximately 150,000 participants from whom T2D cases in this study were ascertained. We genotyped participants using a custom array designed for the Japanese population (Japonica Array v2 [45] or NEO [46]).

#### G-FORCE Study

G-FORCE is a multi-center collaborative study led by the National Hospital Organization, investigating genetic factors related to obesity and T2D in the Japanese population. We genotyped participants using the Infinium Asian Screening Array-24 v1.0 (Illumina).

### Testing dataset

#### Fukushima Diabetes, Endocrinology, and Metabolism (DEM) cohort

The Fukushima DEM cohort is a prospective study at Fukushima Medical University focusing on diabetes and its complications [20,47]. We genotyped participants using the Japonica Array NEO (ToMMo).

### Genotyping QC, phasing, and imputation

For all datasets, we applied an identical quality control (QC) pipeline at both the variant and individual levels. Variants were excluded if they had a call rate <99%, Hardy–Weinberg equilibrium p-value <1.0×10⁻L, or minor allele frequency (MAF) <0.01. Individual-level QC removed samples with call rate <95%, excessive heterozygosity, or prior quality flags (e.g., withdrawal of consent). We performed variant- and sample-level filtering and estimated cryptic relatedness using PLINK v1.9 [48] and v2.0 [49]. We pre-phased genotypes with SHAPEIT2 and imputed them using IMPUTE4, with a combined reference panel from the 1000 Genomes Project Phase 3 v5 (n=2,504) and 3.5KJPNv2 Japanese WGS data (n=3,552). To limit cryptic relatedness, one individual from each pair with PI_HAT >0.1875 was excluded across training and testing datasets.

### Definition of T2D and Control in the TMM, G-FORCE study, and Fukushima DEM cohort

Patients are screened for T2D based on the diagnostic criteria recommended by the World Health Organization (WHO) [50], which classify individuals according to their plasma glucose levels. A diagnosis of T2D is made if an individual meets any of the following conditions: a previous diagnosis of diabetes, a fasting plasma glucose level ≥ 126 mg/dL, a non-fasting plasma glucose level ≥ 200 mg/dL, or an HbA1c level ≥ 6.5%. Individuals under the age of 18 were excluded from the study. In the TMM cohort, participants who self-reported a diagnosis of type 1 diabetes (T1D) were excluded. In the G-FORCE Study and Fukushima DEM cohort, individuals positive for glutamic acid decarboxylase (GAD) antibodies, a key autoimmune marker of T1D, were excluded to avoid inclusion of T1D cases. Non-diabetic controls from TMM were selected by matching on age to achieve balance with the overall T2D case group.

### Subtype assignment by random forest classification

Large cohorts often lack fasting C-peptide/insulin required for computing homeostasis model assessment estimates of beta-cell function (HOMA2-B) and insulin resistance (HOMA2-IR) [51]. In TMM these measures are unavailable, and G-FORCE records only limited clinical items (e.g., approximate age at diagnosis). Therefore, we assigned subtype membership probabilities using a random forest classifier [35], trained on the Fukushima CKD+DEM cohort [20,47]. Specifically, the probability of an individual *i* belonging to a specific subtype *k*, denoted as *P*(*k|***x***_i_*), is computed using the following equation:

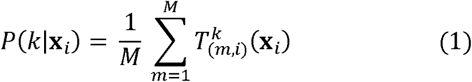

where *i* indexes individuals (1*,…, n*), *m* indexes trees (1*,…, M*), and 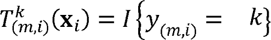 with *I*(.) the indicator function that equals 1 if tree *m* assigns individual *i* to subtype *k*, and 0 otherwise. *y_(m,i)_* denotes the predicted subtype label for individual *i* produced by tree *m* within the forest. Here, *k* is one of the four predefined subtypes x*_i_* denotes the at this step. Let *K* be the number of subtypes. For each individual *i* the RF classifier outputs a vector of baseline clinical covariates supplied to the RF classifier; no genetic predictors are used vector of probabilities over the four subtypes:*{P*(*k|***x***_i_*)*}_k=_*_1*,…k*_, where each element represents the probability of belonging to subtype *k* and the sum of all elements equals one (*∑_k_ P*(*k|***x***_i_*) *=* 1). These subtype membership probabilities were directly utilized in subsequent modeling to perform soft weighting, allowing all individuals to contribute to each subtype-optimized model according to their degree of subtype membership.

### pPS construction

We utilized 12 genetically defined, multi-ancestry T2D variant clusters [27]. These clusters were derived using bNMF, a soft clustering method applied to a variant–trait association matrix comprising 650 T2D-associated variants and 110 T2D-relevant traits. Importantly, individuals in our external validation dataset (Fukushima DEM cohort), who are of Japanese ancestry, were not included in the GWAS summary statistics used to construct this matrix, thereby eliminating any risk of data leakage during model evaluation. For each participant, we computed pPSs as the weighted sum of risk allele dosages across all 650 variants, applying the cluster-specific weights in PLINK v2.0 [49]. Unlike previous studies that applied inclusion thresholds (variant-level cluster weight >0.7802) [27], we did not impose such a threshold and instead included all variants to maximize predictive accuracy.

### Palette PRS construction

The palette PRS is a subtype-optimized polygenic prediction framework that integrates 12 pPSs with subtype-specific pathway weights. To account for uncertainty in subtype classification, we incorporate the subtype membership probability *P*(*k|***x***_i_*) into the loss function.

Specifically, rather than using *P*(*k|***x***_i_*) directly, we define a threshold sigmoid-based weight such that samples with *P*(*k|***x***_i_*) < 0.5 receive a weight of zero and do not contribute to the estimation for subtype *k*, whereas samples with *P*(*k|***x***_i_*) ≥ 0.5 are smoothly weighted according to

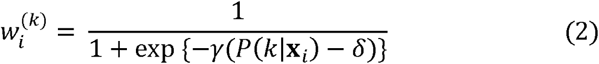

where *γ* (steepness) and δ (threshold) are tuned by grid search with cross-validation.

For each subtype *k*, coefficients are estimated via Elastic-Net–regularized binomial logistic regression to prevent overfitting and ensure robust estimation. Let *y_i_* be binary if the individual belongs to subtype *k* then 1, other 0. Given *w_i_* and *y_i_*, the optimization problem is

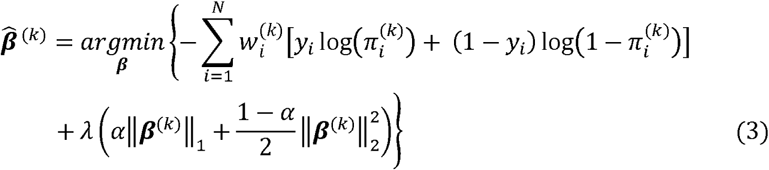

where 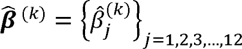 is a coefficient vector specific to subtype *k*, and 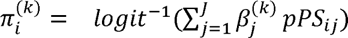. Each element 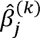 corresponds to pathway *i* and *j* is the number of pathways. *α* ∈ [0,1] is a hyperparameter to control the L1/L2 trade-off; and *λ* ≥ 0 sets overall regularization strength. *λ* and *α* are selected by cross-validation as well as *γ* and *δ*. The subtype-optimized palette PRS for individual *i* is

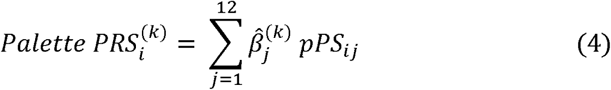

where 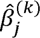 is the coefficient for pathway *j* in subtype *k*, and *pPS_ij_* is the partitioned polygenic risk score for individual *i*. This design integrates pathway and subtype information while down-weighting uncertain subtype labels via the sigmoid weights, thereby yielding robust and flexible subtype-optimized risk prediction.

### Stratified GWAS-based PRS and Overall T2D PRS (650 variants)

As benchmarks for comparison with our proposed palette PRS, we first constructed subtype-optimized PRS using summary statistics from the largest T2D subtype-stratified GWASs [21] (Supplementary Fig.5). These PRSs were generated using PRS-CS-auto [37], which applies a continuous shrinkage prior to effect sizes while accounting for linkage disequilibrium. These subtype-optimized PRS served as alternative benchmarks, enabling comparison between GWAS-derived subtype-optimized approaches and our pathway-based palette approach. In addition, we constructed an overall T2D polygenic risk score (PRS) based on 650 genome-wide significant variants reported by Smith et al. [27] (Supplementary Fig.5). Risk allele dosages were weighted by the corresponding β coefficients from the MVP+DIAMANTE multi-ancestry GWAS summary statistics [36]. Overall T2D PRS as a single homogeneous phenotype without accounting for clinical heterogeneity and thus serves as a reference for evaluating the added predictive value of the subtype-specific palette PRS. The overall T2D PRS ignores clinical heterogeneity and thus serves as a baseline for evaluating the added value of the subtype-specific palette PRS.

### Software and statistical computing

All statistical analyses were performed using R (version 4) and Python (version 3). Genome-wide data processing and quality control were conducted using PLINK v1.9 [48] and v2.0 [49].

## Ethics declarations Conflicts of interest

The authors declare that they have no conflict of interest.

## Ethical standard statement

This multi-center study was centrally approved by the Institutional Ethics Review Board of the Tohoku University Tohoku Medical Megabank Organization, and the protocol was conducted in accordance with the Declaration of Helsinki.

## Informed consent

All subjects gave written informed consent for their participation in the study and for genetic analysis.

## Data availability

Individual-level data from Japanese cohorts are not publicly available due to ethical restrictions. The UKBB analysis was conducted under application number 76615 (https://www.ukbiobank.ac.uk). GWAS summary statistics used in the calculation of the T2D PRS were obtained from dbGaP under accession number phs001672.v3.p1 (Veterans Administration Million Veteran Program Summary Results from Omics Studies). Subtype-stratified GWAS summary statistics from the study by Mansour Aly et al [21] are publicly available in the GWAS Catalog (www.ebi.ac.uk/gwas/) under accession numbers GCST90026413–GCST90026417. Data from the NHANES III cohort is also publicly available at (https://wwwn.cdc.gov/nchs/nhanes/Default.aspx)

## Code availability

The code used for constructing the palette PRS, including subtype-specific weighting and model training procedures, will be made publicly available upon publication of this article.

## Acknowledgements

This work was supported by JSPS KAKENHI (Grant Number JP24K13523), the JST Moonshot R&D Program (Grant Number JPMJMS2023), the Tohoku Medical Megabank Project at Tohoku University (Grant Number JP21tm0124005), AMED (Grant Number JP22tm0424224), and RIKEN AIP through the subsidy for the Advanced Integrated Intelligence Platform project of MEXT, Japan. We are deeply grateful to all participants and investigators of the TMM, Fukushima DEM, CKD cohort, G-FORCE study, and the UK Biobank (UKB). We would also like to acknowledge everyone who assisted with the study, especially Hideki Katagiri, Sachiyo Sugimoto, Aye Ko Ko Minn, Kazuki Kumada, and Atsushi Hasegawa. We also thank ToMMo for providing the supercomputing resources used in this study and appreciate the illustration service from MEDICAL FIG. (Medical Education Inc.).

## Author contributions

G.T. designed the study and supervised the project. A.M., H.T., A.N., R.S., T.O., T.K., and contributed to the study design. A.M. and H.T. performed the analyses. C.G. contributed to data analyses. H.T. and M.S. contributed to the management of data from the Fukushima DEM cohort. H.Y. and T.A. contributed to the management of data from the G-FORCE cohort. K.T. and J.K. contributed to the management of data from the Fukushima CKD study. A.M. wrote the manuscript with critical input from H.T., A.N., R.S., T.O., J.T., M.S., and G.T.

## Notes

### Competing Interest Statement

The authors have declared no competing interest.

