## Supplementary Files for "Palette polygenic risk score framework improves risk prediction by capturing clinical heterogeneity of type 2 diabetes"

**Supplementary Figure legends**

**Supplementary Fig.1 Distribution of each subtype and their clinical characteristics across cohorts.** Pie charts show the relative proportions of T2D subtypes (SIDD, SIRD, MOD, MARD, and Undecidable) identified in each cohort. Boxplots display the distributions of age at diagnosis, BMI, HbA1c, HOMA2-B, and HOMA2-IR for each subtype within the corresponding cohort. These clinical profiles illustrate the characteristic differences among subtypes and the consistency of subtype patterns across populations.

**Supplementary Fig.2 Association of pPSs with T2D subtype.** Association of pPSs with T2D subtypes evaluated using univariate logistic regression models for (a) the Japanese population (SIDD, N=320; SIRD, N=308; MOD, N=1,106; MARD, N=4,542; Control from the training dataset, N=20,525) and (b) UK Biobank British participants (SIDD, N=1,563; SIRD, N=508; MOD, N=3,504; MARD, N=4,441; Control, N=413,815). Odds ratios per 1-standard deviation increase in each pPS with 95% confidence intervals, using control individuals as the reference group.

**Supplementary Fig.3 Stratified distributions of partitioned polygenic scores (pPS) by T2D subtypes.** Kernel density distributions of normalized pPS are shown across T2D subtypes. Dots on each density curve indicate the median. 12 pPSs are presented for two study populations: (a) the Japanese cohort, comprising SIDD (N=320), SIRD (N=308), MOD (N=1,106), MARD (N=4,543), and controls (N=20,525); and (b) the UK Biobank British participants, comprising SIDD (N=1,563), SIRD (N=508), MOD (N=3,504), MARD (N=4,441), and controls (N=413,815).

**Supplementary Fig.4 Calibration plots of subtype-specific palette PRS models.** Calibration performance of the palette PRS is shown separately for each T2D subtype. The x-axis represents the mean predicted risk, and the y-axis represents the mean observed risk. The blue line indicates the locally estimated regression (LOESS) fit with 95% confidence intervals (shaded area), and error bars denote binomial confidence intervals of observed proportions. The dashed diagonal line indicates perfect calibration.

**Supplementary Fig.5 Stratified GWAS-based PRS and overall T2D PRS (650 variants).**

Subtype-specific PRS were constructed from the largest subtype-stratified GWAS of T2D using PRS-CS-auto, and an overall T2D PRS was built from 650 genome-wide significant variants. Predictive performance of these GWAS-derived scores was evaluated in the Fukushima DEM cohort using AUC.

**Supplementary Fig.6 Subtype classification and model construction in the UK Biobank.** British participants in the UK Biobank were classified into subtype using the Random Forest classifier trained on NHANES Ⅲ.

**Supplementary Fig.7 Training dataset derived from NHANES III.** The training dataset from NHANES Ⅲ was generated by applying k-means clustering (k=4) to individuals with T2D, based on age at diagnosis, BMI, HbA1c, HOMA2-B, and HOMA2-IR.

**Supplementary Fig.1**

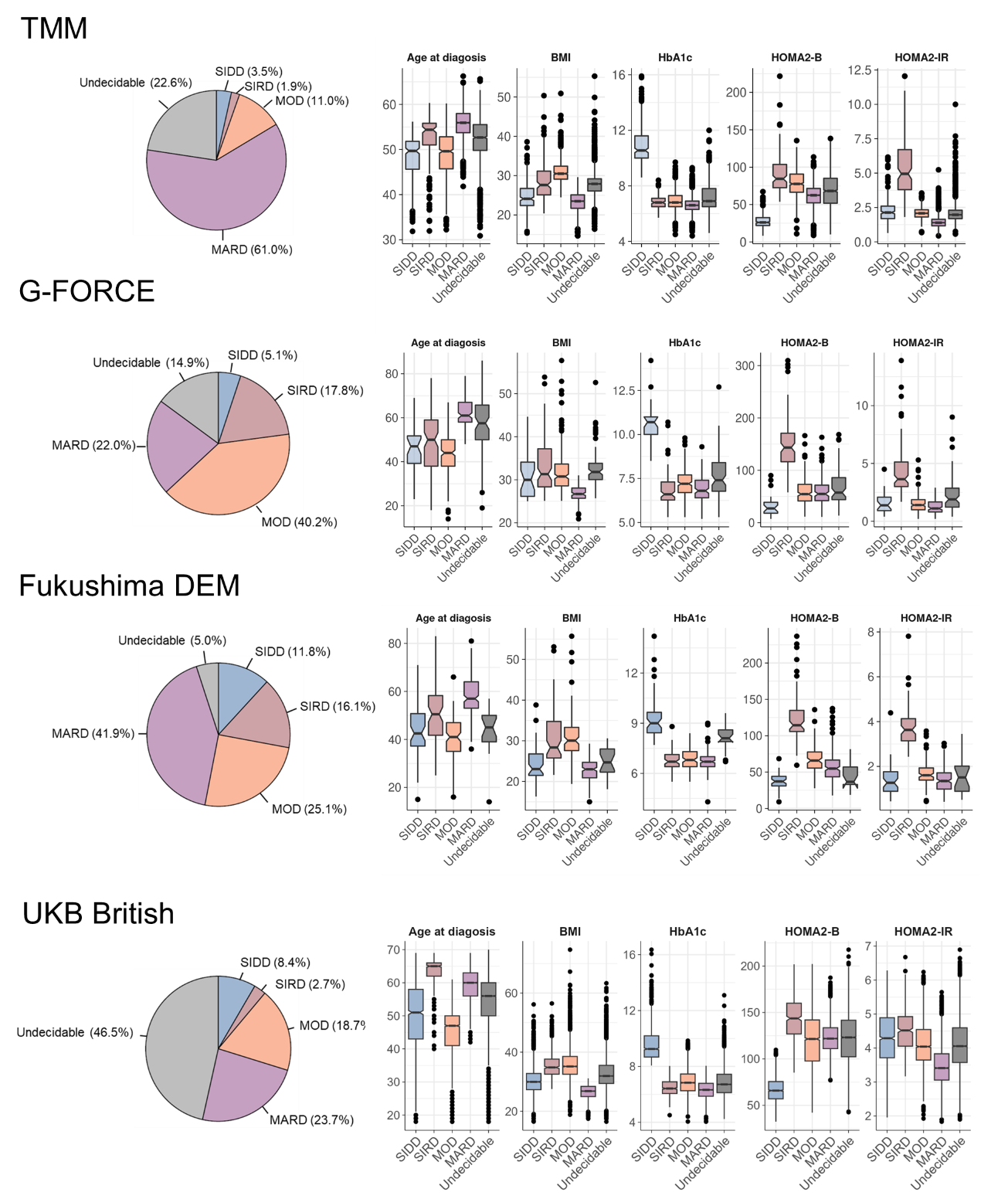

**Supplementary Fig.2**

a. **Japanese population (TMM, G-FORCE, and Fukushima DEM)**

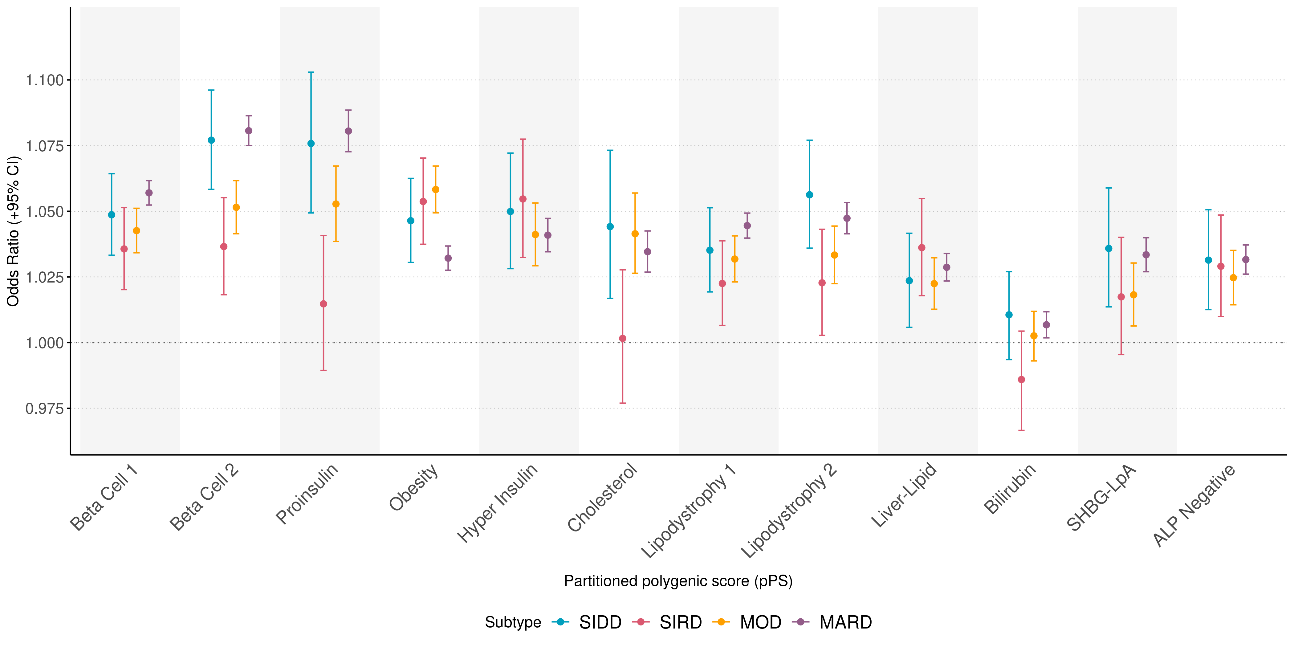

b. **UKB British**

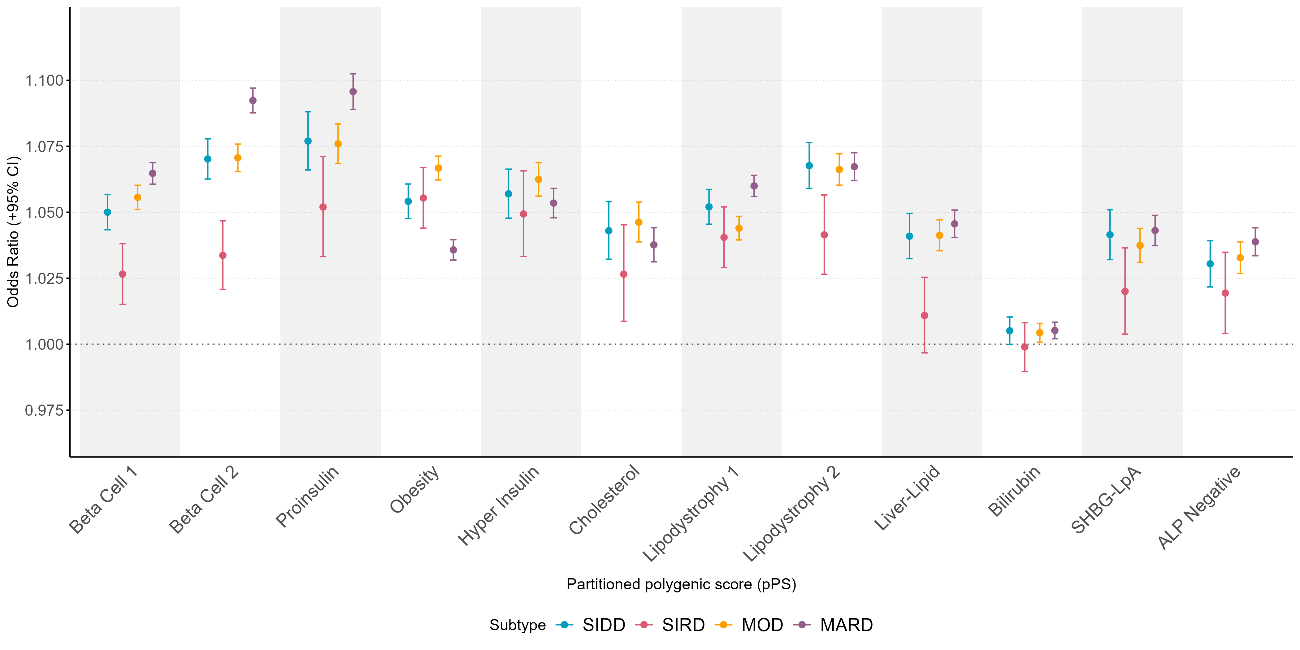

**Supplementary Fig.3**

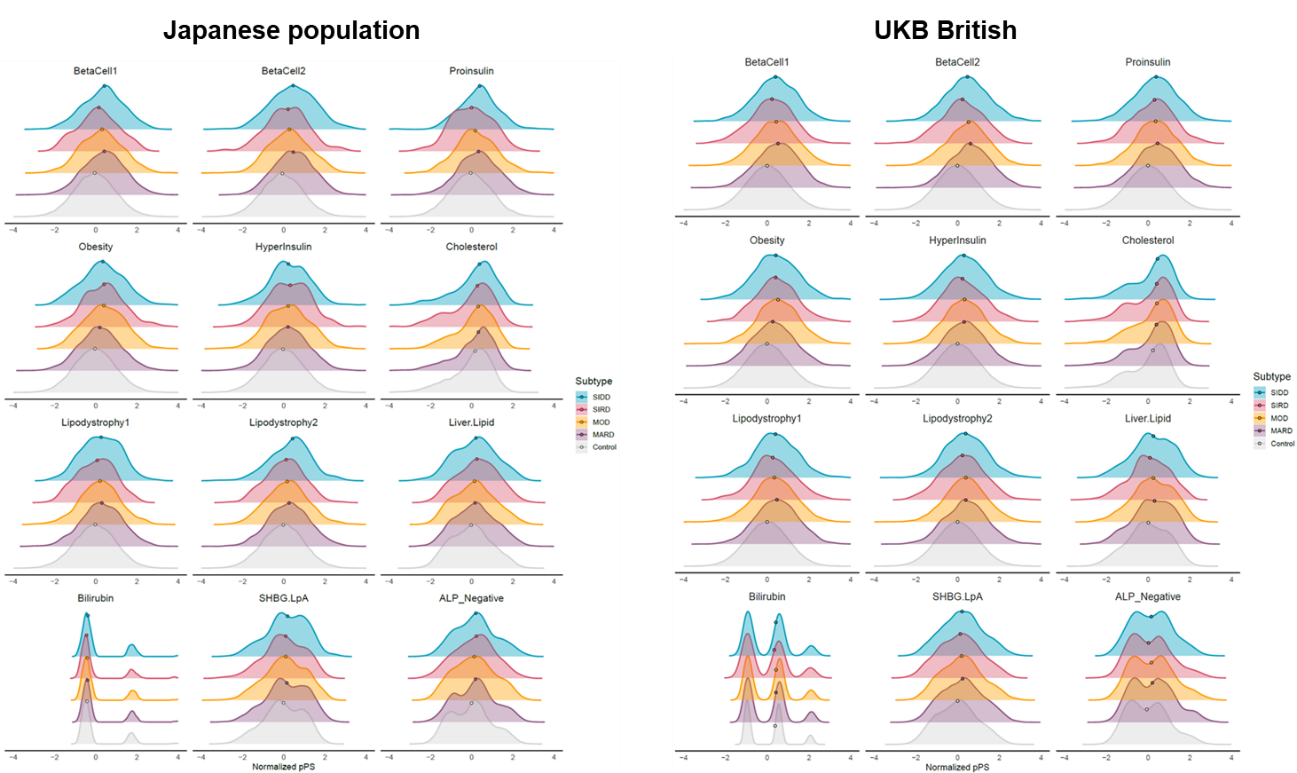

**Supplementary Fig.4**

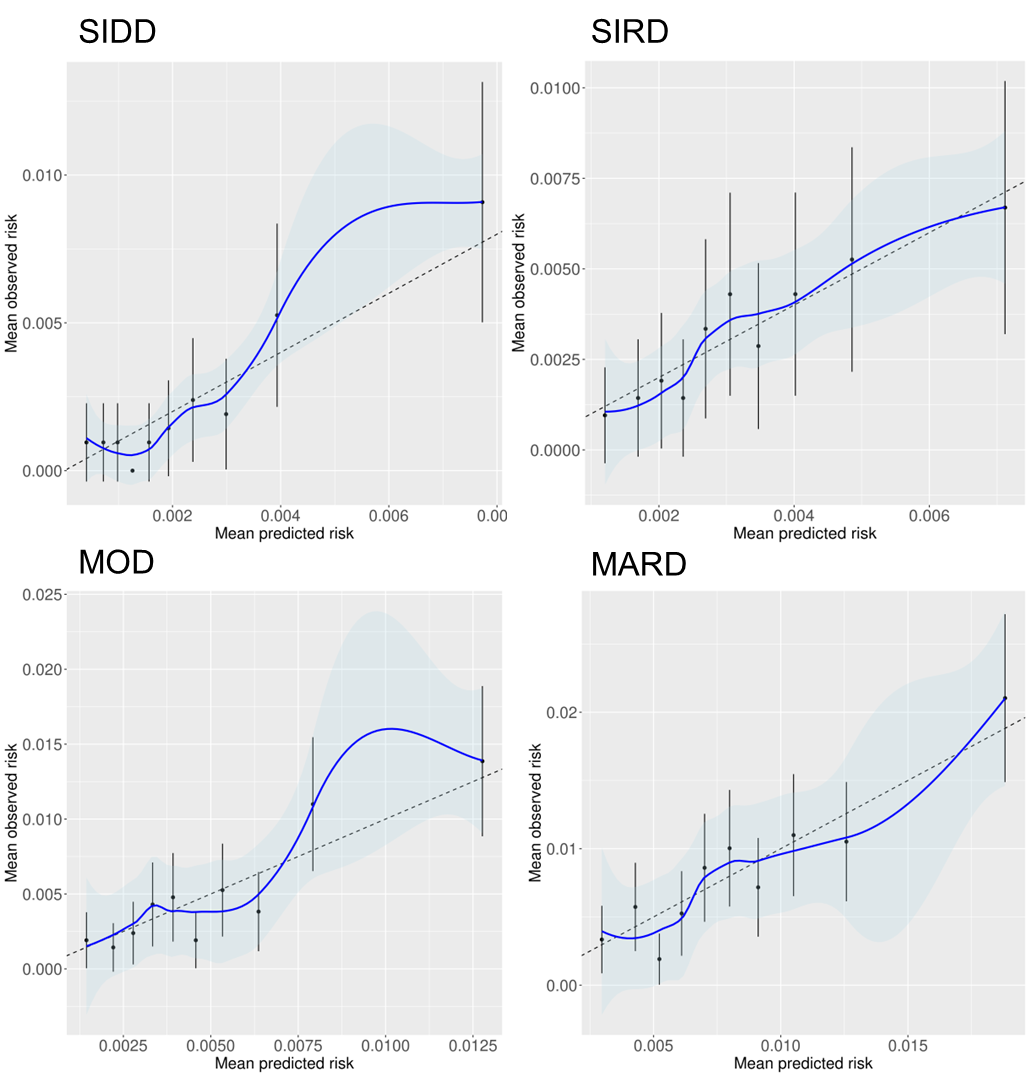

**Supplementary Fig.5**

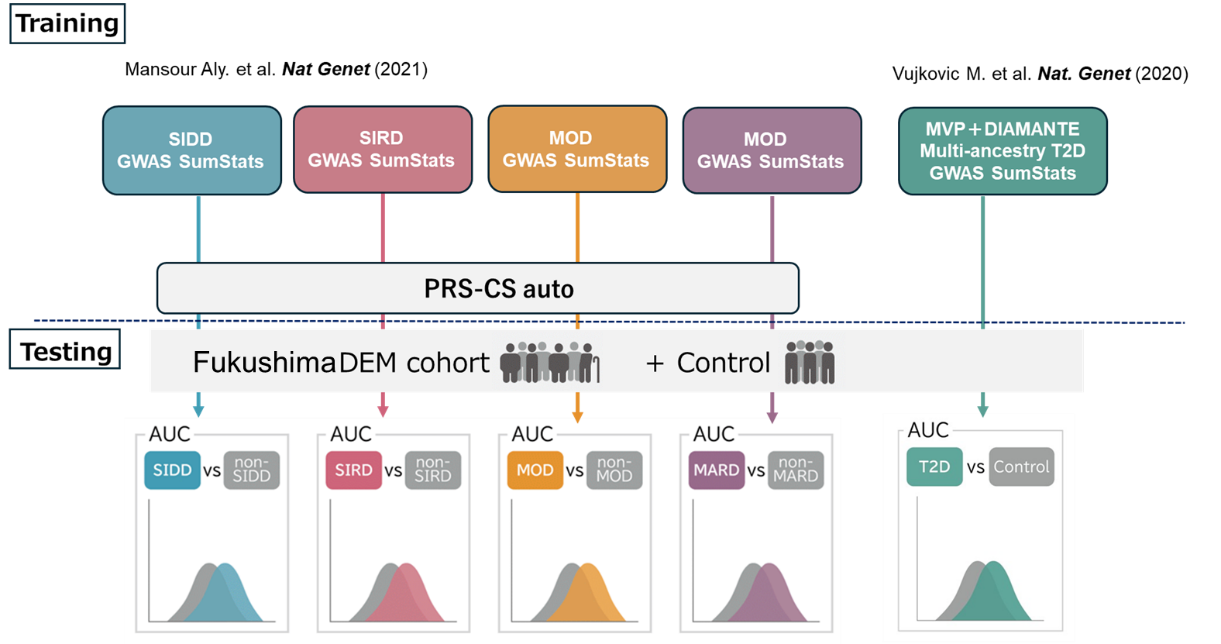

**Supplementary Fig.6**

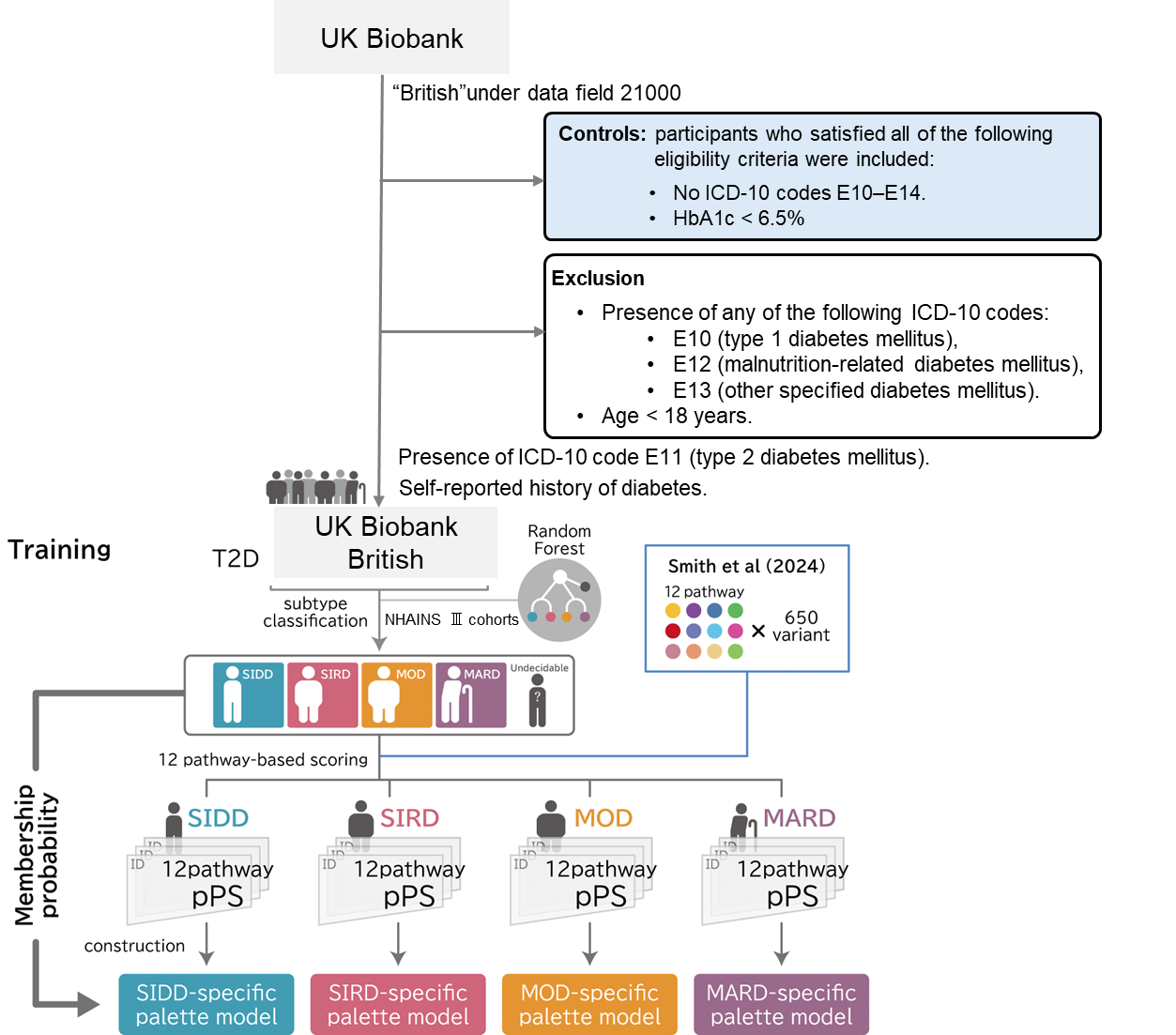

**Supplementary Fig.7**

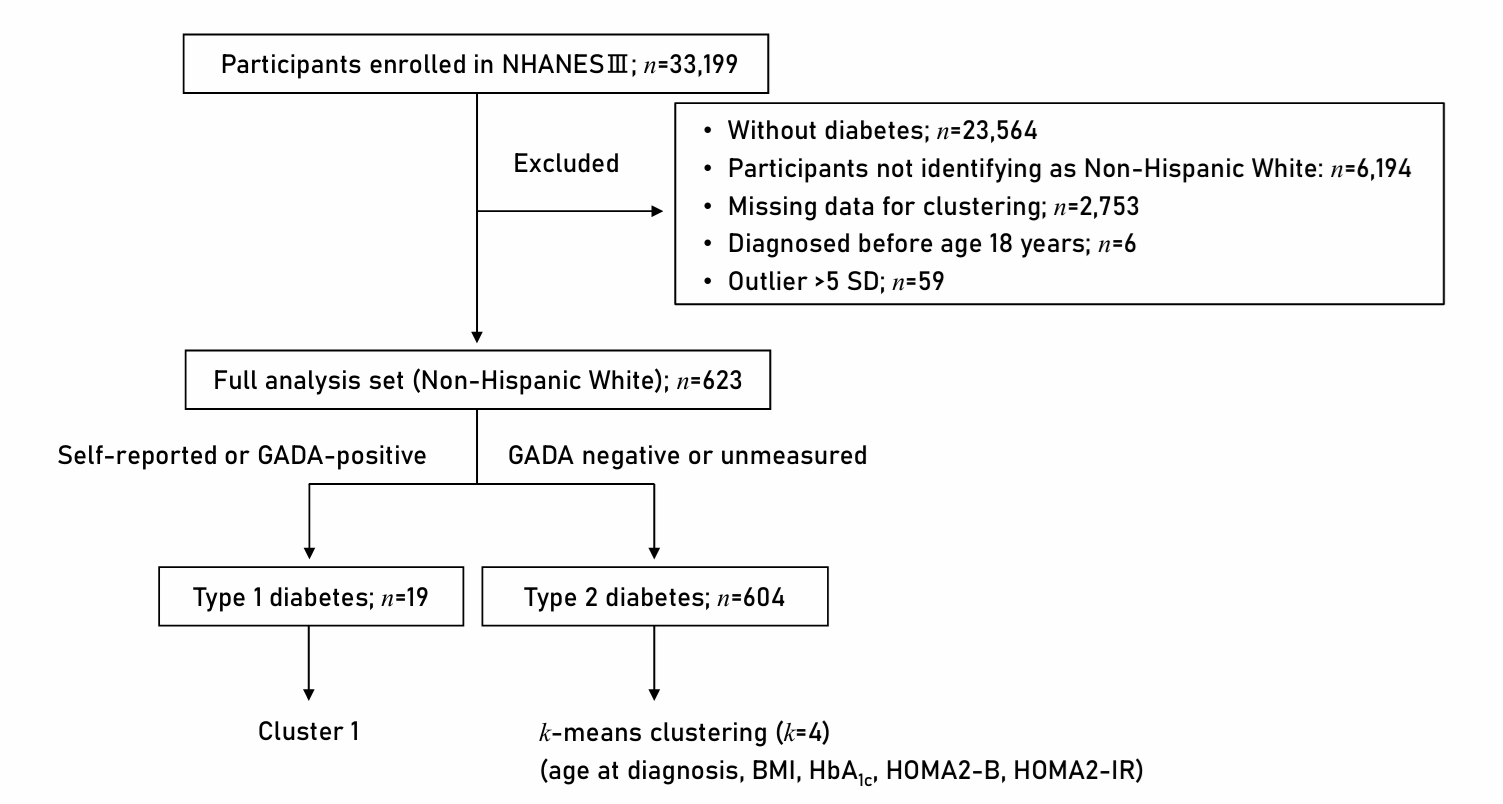

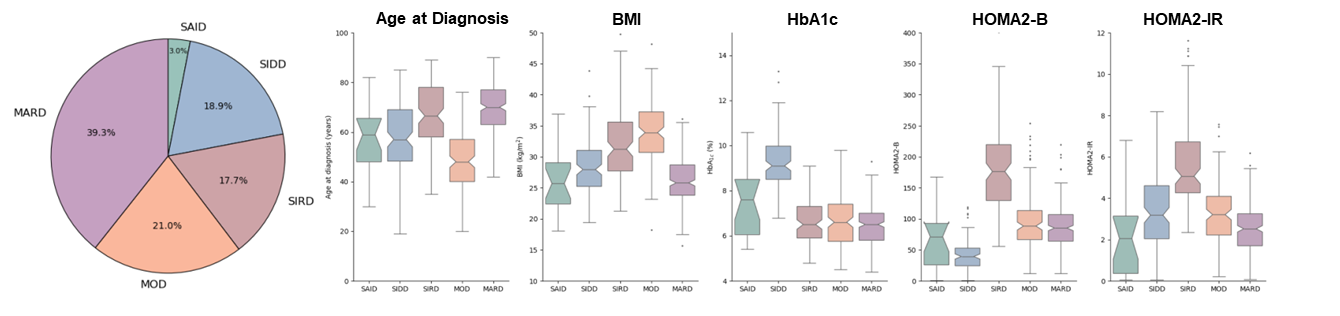
